# Survey of SARS-CoV-2 genetic diversity in two major Brazilian cities using a fast and affordable Sanger sequencing strategy

**DOI:** 10.1101/2021.07.02.21259802

**Authors:** Erick Gustavo Dorlass, Karine Lima Lourenço, Rubens Daniel Miserani Magalhães, Hugo Sato, Alex Fiorini, Renata Peixoto, Helena Perez Coelho, Bruna Larotonda Telezynski, Guilherme Pereira Scagion, Tatiana Ometto, Luciano Matsumiya Thomazelli, Danielle Bruna Leal Oliveira, Ana Paula Fernandes, Edison Luiz Durigon, Flavio Guimarães Fonseca, Santuza Maria Ribeiro Teixeira

## Abstract

Genetic variants of SARS-CoV-2 have been emerging and circulating in many places across the world. Rapid detection of these variants is essential since their dissemination can impact transmission rates, diagnostic procedures, disease severity, response to vaccines or patient management. Sanger sequencing has been used as the preferred approach for variant detection among circulating human immunodeficiency and measles virus genotypes. Using primers to amplify a fragment of the SARS-CoV-2 genome encoding part of the Spike protein, we showed that Sanger sequencing allowed us to rapidly detect the introduction and spread of three distinct SARS-CoV-2 variants in two major Brazilian cities. In both cities, after the predominance of variants closely related to the virus first identified in China, the emergence of the P.2 variant was quickly followed by the identification of the P1 variant, which became dominant in less than one month after it was first detected.

## 1. Introduction

After the announcement of the SARS-CoV-2 pandemic in March of 2020, different virus variants have emerged and spread fast, many of them becoming dominant over previous ones. These variants have been categorized as Variants of Interest (VOI) or Variants of Concern (VOC) [1]. Although the appearance of new variants is expected, as SARS-CoV2 continues to evolve and differentiate in response to different host-derived selective pressures, only four of the currently identified variants are considered as VOC: B.1.1.77 (UK/Alpha variant), P.1 (Brazil/Gamma variant), B.1.351 (South Africa/Beta variant) and B.1.617.2 (India/Delta variant). These four variants have specific amino acid substitutions in the Spike protein, two of them known to be associated with increased infection and mortality: E484K and N501Y [2–4]. Also, some studies have shown a decreased in the immune response when convalescent serum and immunized serum from vaccinated individuals were tested against variants with these mutations [5–8].

The P.1 variant, which is thought to have evolved from the B.1.1.28 lineage in a period of fast molecular evolution dated around mid-November of 2020, is of particular interest in this study [9]. The P.1 emergence has been unequivocally traced to the city of Manaus, and it is thought to have been one of the main factors responsible for the upsurge of COVID-19 cases and deaths in this city during December 2021/January 2021 period, leading to the complete collapse of the local health system. The P.1 genetic background includes 21□missense, lineage-specific mutations, 10 of which are located within the Spike protein (L18F, T20N, P26S, D138Y, R190S, K417T, E484K, N501Y, H655Y and T1027I) [10]. Circulating undetected at least until mid-February 2021, the mutant rapidly spread to other Brazilian cities, most likely due to infections carried by people fleeing the collapsed Amazon city. Different from P.1, the P.2 variant bears the E484K mutation only, but not the other two mutations of concern, N501Y and K417T. Four other lineage-defining mutations were also found in other parts of the genome, including UTR, Orf1ab and N. Genomic surveillance of SARS-CoV-2 is an essential component of the epidemiological profile of the COVID-19 pandemic since variant identification can be associated with new outbreaks and re-infection. Fast and precise detection of these variants is crucial for public health officials to monitor the impact caused by their introduction in the population of circulating viruses so that control measures can be adjusted accordingly. The high cost of complete genomic sequencing still prevents low/medium-income countries to properly perform such surveillance using Next-Generation Sequencing (NGS) platforms. The more traditional Sanger Sequencing may be an alternative for identification of circulating variants since it can provide results very rapidly and be performed using a sequencing infra-structure already in place in many laboratories. Using Sanger sequencing, we identified circulating VOI and VOCs from samples collected in two major cities in Brazil during a period of 12 months. São Paulo, with an estimated population of 12,325,232 [13], and Belo Horizonte, with 2,521,564 citizens [14] are the first and sixth largest cities in Brazil and have reported a total of 1,048,061 [15] and 177,432 [16] cases of COVID-19 as of April 30.

## 2. Methods

### 2.1. Ethical Statement

Part of study involving samples from São Paulo city was approved by Instituto de Ciências Biomédicas ethical committee. CAAE number: 32712620.9.0000.5467. The study involving samples from Belo Horizonte was approved by the UFMG Ethics Committee, CAAE-35074720.3.0000.5149.

### 2.2. Sample collection and SARS-CoV-2 identification

Nasopharyngeal swabs from patients with airway infection symptoms were collected from seven hospitals in the city of São Paulo, SP as well as from a specialized COVID-19 Health Center (UPA Centro-Sul) in Belo Horizonte, MG, Brazil. Part of the samples from the Belo Horizonte metropolitan area, which includes the cities of Betim, Contagem, Ribeirão das Neves e Santa Luzia, were obtained from Fundação Ezequiel Dias (Funed-MG). RNA was extracted either with Viral Pathogens kit (Thermofisher) in MagMax or with the QIAamp Viral RNA Mini Kit (Qiagen, USA), according to protocols provided by the manufacturers. RT-qPCR was performed using primers and probes described in the Berlin (Charité/Berlin) protocol [17], targeting the gene E from SARS-CoV-2 and the human RNAse P mRNA, used as endogenous amplification reaction control. All samples tested positive were selected for gene amplification with PCR primers targeting the Spike gene and sequencing using the Sanger method.

### 2.3. Primer selection and PCR amplification

Primers specific for the amplification of a 1006 bp region, codifying part of subunit one of the Spike protein of SARS-CoV-2, was selected in the ARTIC protocol [18]. The primers are shown in **Supplementary table**. The 1006 bp amplified region encodes amino acids K417, L452, T478, E484, N501 and A570, which have been associated with the characterization of several VOI and the VOCs. The same pair of primers together with a third primer were used for all sequencing reactions. PCR amplifications were performed after Reverse Transcription reactions using random primers and the High-Capacity cDNA Reverse Transcription Kit (thermofisher) following manufacturer’s instructions. PCR amplifications were performed with the Invitrogen Platinum Taq DNA Polymerase kit and the following amplification protocol: 5 min at 95°C followed by 40 cycles of 30 sec at 94°C, 30 sec at 56°C and 80 sec at 72°C with a final extension time of 5 min at 72°C. The amplicons were analyzed by 1% agarose gel electrophoresis. After treatment with Exosap-it (Thermofisher), sequencing reactions were performed with the BigDyeTM Terminator v3.1 Cycle Sequencing Kit (Applied Biosystems) and either the ABI 3730 or the ABI PRISM 3130XL Genetic Analyzer according to manufacturer’s instructions.

### 2.4. Sequence Analyses

Assembling of Sanger contigs were performed with the GeneStudio software [19]. Each primer corresponding sequence was trimmed in a fixed position (40 and 450) followed by manual evaluation of aligned chromatograms for each sample. By this approach 228 contigs were constructed. These sequences were aligned by the MAFFT tool [20,21] with default parameters: 0 “Offset”, 1.53 “gap open penalty”, 0 “Maximum number of iterative refinements”. Benchling platform[22] and AliView software [23] were used for visualization of alignments. The samples were manually genotyped by evaluation of the modifications of interest in the contigs (K417N/T, L452R, T478K, E484K, N501Y, A570D), after in silico translation of the consensus nucleotide sequences using standard genetic code. Stacked Bar Plots were constructed with the R ggplot2 package.

Next-Generation Sequencing (NGS) was performed with 16 samples using the Ion Torrent Platform with SARS-CoV-2 Ion Ampliseq panel. Sample libraries were constructed on the Ion Chef system with the Ion Ampliseq Chef DL8 kit. Sequencing was performed in two 530 Ion Chip with a capacity of 20 million reads of each chip. Sequencing analysis was performed on Torrent server with IRMA plugin for read assembly and SnpEff for variant annotation. All final sequences were submitted to Panglolin [24] for lineage assignment.

To construct the dendrograms, the aligned Sanger contigs were trimmed in 5’ and 3’ end to eliminate all gaps and divergent sequencing quality. Nine short contigs (PAC1346B, COV958, HIDV3706, PAC1705, PAC1803, ECO207, PAC721, COV701, HIDV4996) were excluded to maximize the size of multiple alignment after trimming. An alignment of 770 nt was obtained after these steps. The genomic sequences were aligned using the MAFTT tool using default parameters. The phylogenetic trees were constructed using the iqtree tool [25], with the “Maximum likelihood” statistical approach and substitution model TPM3+F+I to Sanger and GTR+F+I NGS dataset. Lineage B (GISAID_ID: EPI_ISL_402123, 2019) was defined as “outgroup” and the R ggtree package [26] was used for the tree visualization.

## 3. Results

A total of 228 SARS-CoV-2 positive samples were successfully amplified using the primers shown in **Figure 1**. The amplicons correspond to viruses present in positive samples from 130 infected individuals from the city of São Paulo collected between March/2020 and Abril/2021 and 98 positive samples from individuals from the Belo Horizonte metropolitan area collected between August/2020 and Abril/2021. As shown in **Table 1**, mutations in the 1006 bp amplified region corresponding to amino acids positions 417, 484, 501 and 570 of the Spike protein allowed the identification of several VOIs and VOCs. Because it was not possible to discern between the P.2 and N.9 variants, based on the sequencing of the 1006 bp amplicon, we also amplified an additional region of 969 bp, containing the V1176F mutation, allowing us to discriminate between these two variants (**Figure 1**). Five samples classified as P.2/N.9 were sequenced and all of them had the V1176F mutation, and was then classified as P.2 (data not shown). Based on the sequences of the amplified 1006 bp fragment generated from all 228 samples collected from individuals in São Paulo and Belo Horizonte, we were able to identify, besides the lineage that is closely related to the original strain isolated in Wuhan (B.1) the B.1.1.7 variant, which have been previously identified in England as well as the P.1 and P.2/N9 variants which have been previously identified in Brazil. **Figure 2** showed that, while in São Paulo city, the P.2/N.9 variant appeared in September 2020 (**Figure 2A, C**), in Belo Horizonte, the same variant was first identified in November (**Figure 2B, D**). The emergence of the P.2 variant was announced in December, after being identified in the state of Rio de Janeiro [11] and rapidly spread throughout the country. Our data showed that this lineage was actually circulating in the neighbor state of São Paulo at least 3 months before this announcement and was already present in Belo Horizonte, also a large city close to Rio de Janeiro, one month before. **Figure 2** also showed that, in the two cities, the P.2 variant was rapidly replaced by the P.1 variant, which was initially described in the city of Manaus, more than 3,000 Km distant from São Paulo and Belo Horizonte. In both cities, the P.1 variant was first detected in February. It is interesting to note that in both cities, one month after its appearance, the P.1 variant became largely predominant among all other circulating lineages, being identified as 75 % and 86 % of all samples analyzed in Belo Horizonte and São Paulo, respectively, in March 2021. Whereas in Belo Horizonte, the P.1 variant corresponded to all samples collected in the month of April, one sample corresponding to the P.2 variant was still identified in April in São Paulo city. It is also noteworthy that, in contrast to the P.2 variant, which dominated over the B.1 lineage, and the P.1 variant, which rapidly dominated over the P.2 variant, the appearance of the B.1.1.77 variant in both cities during the month of March did not result in a similar, fast dissemination. Because the B.1.1.77 variant was introduced in both cities after the P.1 variant was already in place, it can be assumed that its dissemination capacity is significantly lower compared to P.1.

**Table 1.** Amino acid changes found in circulating variants that can be identified by sequencing the amplified fragments.

| Variants | Mutations in the amplified fragment |
| --- | --- |
| P.1 (VOC) | K417T, E484K, N501Y |
| P.2 (VOI) | E484K |
| B.1.1.7 (VOC) | N501Y, A570D |
| B.1.351 (VOC) | K417N, E484K, N501Y |
| B.1.617.2 (VOC) | L452R, T478K |

**Figure 1.**
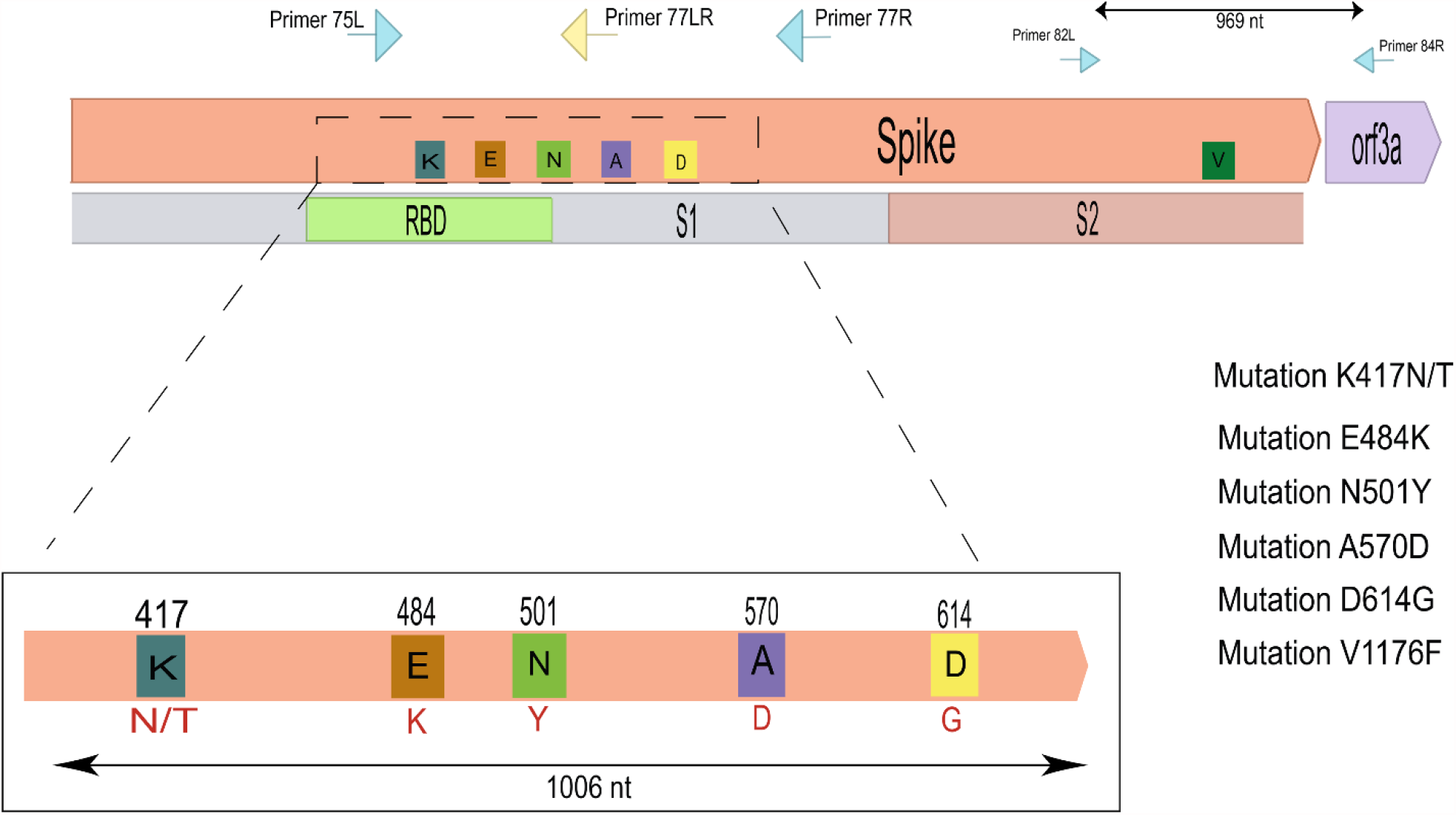
Schematic representations of the Spike protein and the 1006 bp amplicon. PCR amplifications were performed with primers that were used both for PCR and Sanger sequencing (blue arrows). Yellow arrows indicate primers used only for sequencing. Colored rectangles represent changes in amino acids found in different VOI and VOC’s and their respective positions in the SARS-CoV-2 genome, Lysine (K) blue, Glutamic acid (E) orange, Asparagine (N) green, Alanine (A) purple, Aspartic acid (D) yellow, Valine (V) dark green. It is also represented the 969 bp PCR amplificon generated with primers annealing within the spike and orf3a protein genes (small blue arrows), which allowed us to verify for the presence or absence of a mutation at position 1176 that differs in the VOIs N.9 and P.2.

**Figure 2.**
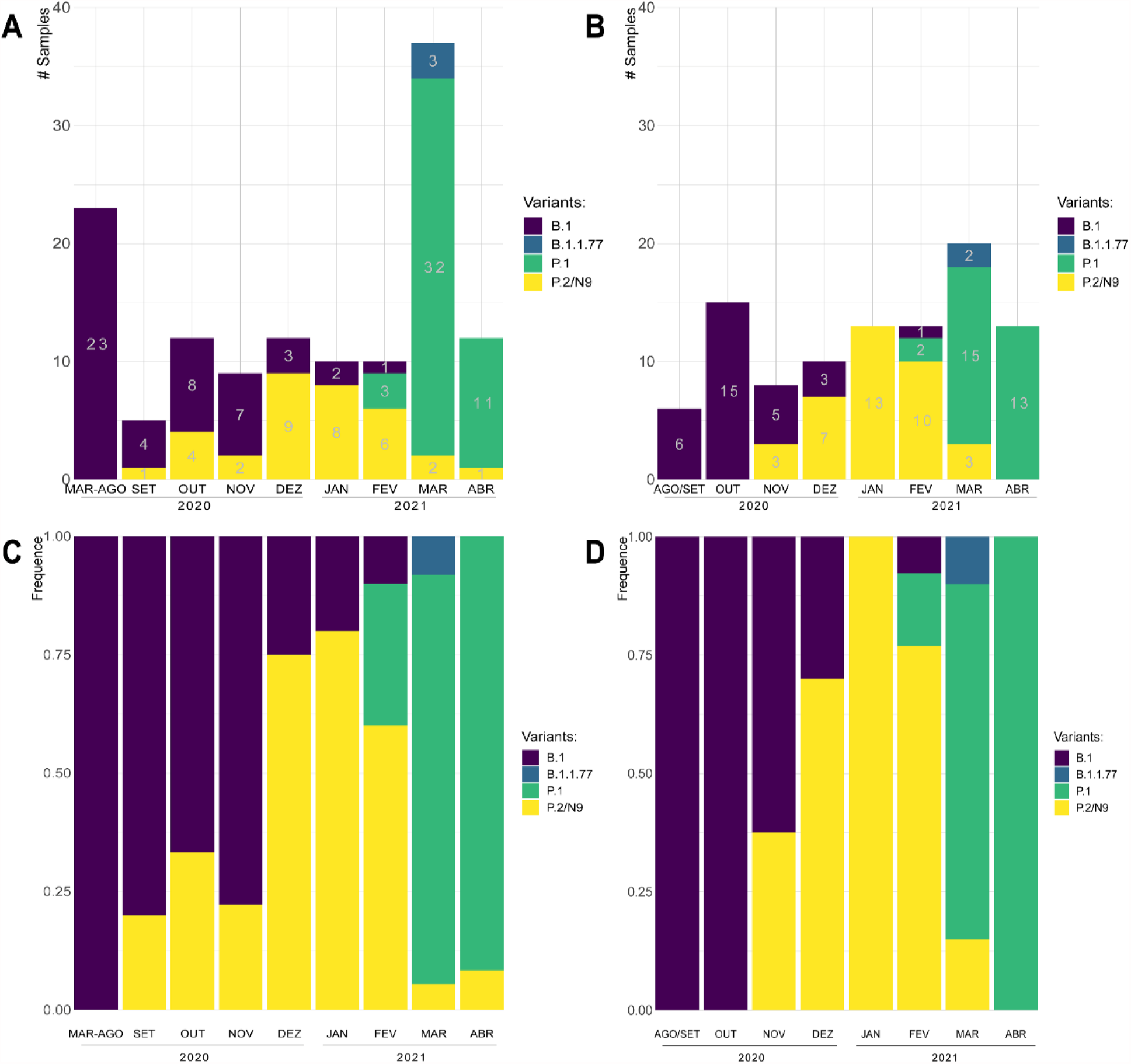
Stacked bar plots of the SARS-CoV-2 variants genotyped by Sanger sequencing in the cities of São Paulo (**A, C**) and in the metropolitan region of Belo Horizonte (**B, D**). Colors of the bars represent the number of samples (**A, B**) or the frequency (**C, D**) of the variants by period: purple, B.1; blue, B.1.1.7; green, P.1; yellow P.2 / N.9.

Phylogenetic and molecular evolution analyses were performed using nucleotide sequences derived from 219 samples. Nine of the 228 samples were not included in this analysis because the contig size became too short after trimming the sequence extremities. Although these sequences also contain mutations of interest, they were excluded from the phylogenetic analyses because the differences in size cause a considerable impact on the multiple alignment. As shown in **Figure 3A**, all generated sequences were grouped in four main clusters corresponding to the B.1 lineage and the B.1.1.7, P.1 and P.2/N.9 variants. This analysis also confirms that the B.1.1.7 was derived from the B.1 lineage, as previously described, and also indicated that the P.1 and P.2/N.9 variants have emerged independently in Brazil.

**Figure 3.**
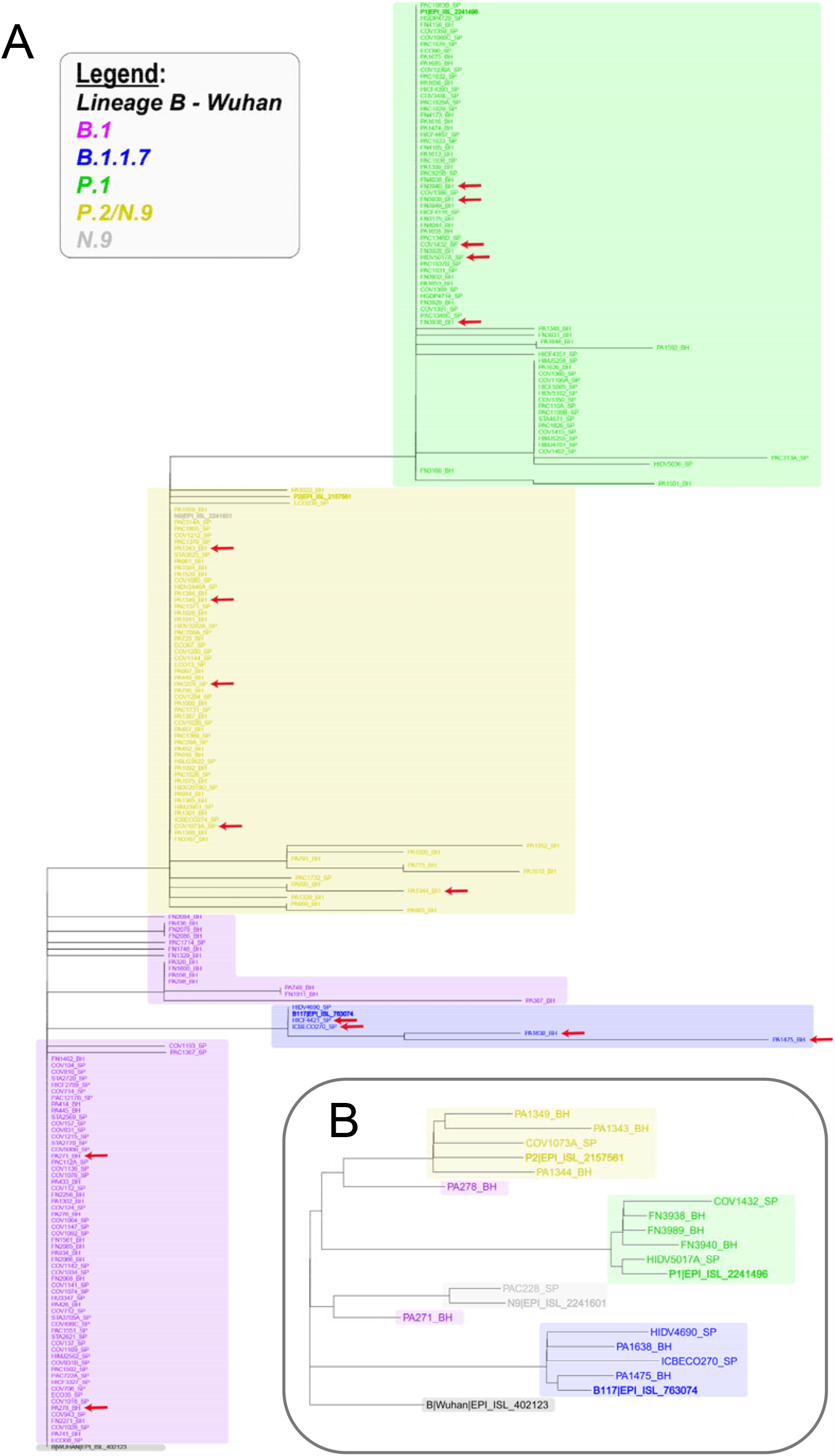
Dendrogram showing the relationship between the sequences obtained by Sanger (**A**) and by NGS sequencing of the complete genome of some of these samples (**B**). Samples whose complete genome has been sequenced are indicated in **A** by the red arrow. The genotyped sequences of each of the variants of interest are represented in colors: purple, B.1; blue, B.1.1.7; green, P.1; yellow, P.2 / N.9. The dendrograms were rooted using the corresponding 770 bp region of Lineage B (Wuhan, EPI_ISL_402123,2019). Reference sequences for each variant of interest are highlighted in bold.

To confirm that the sequence obtained from 1006 bp fragment of the Spike gene was sufficient to unambiguously ascertain the identity of the variants, we selected 16 samples to determine their complete genome sequences using the Ion Torrent Sequencing Platform. The viral samples that have their complete genomes sequenced were indicated by an arrow on **Figure 3A**. The phylogenetic tree shown in **Figure 3B** indicates that, for all genomes that were fully sequenced, the classification based on partial Sanger sequencing as belonging to B.1 lineage and the B.1.1.7, P.1 and P.2/N.9 variants could be fully validated by the complete NGS genome analyses.

Using Sanger sequencing, we have also analyzed two samples that were collected during the third week of May 2021, at the international airport of Belo Horizonte. These samples were from RT-PCR-positive individuals who had contact with a flight crew with Indian origin. In less than 5 days, we were able to amplify the 1006 bp fragment, sequence and verify that in both cases, they correspond to the P.1 variant, and not to the new emerging variant B.1.617.2, which has the two distinct mutations in this region. Taken together, these results showed the power of the Sanger sequencing strategy to rapidly generate data that can be used to perform genomic surveillance during the COVID-19 pandemic. It also confirmed that it can be employed for genomic epidemiology studies of other diseases. With a relatively small number of samples, we showed that the introduction of VOI or VOCs in two large urban areas can rapidly reshape the genomic profile, which may have a significant impact in the course of the COVID-19 pandemic.

## 4. Discussion

In the first months of the SARS-CoV2 pandemics it became clear that ample qPCR testing and contact tracing would be essential to mitigate local and nationwide virus spread, limiting overall impacts of the disease [27,28]. Indeed, countries that exceeded in testing their population had relative success in diminishing the burden of the epidemics, both in terms of number of affected people as well as in social and economic impacts [29,30]. Nonetheless, over and over the new coronavirus has proven to be an elusive enemy to defeat. One of the most difficult challenges related to the current moment of the pandemic is certainly to identify and contain the spread of virus variants presenting mutations that confer dangerous selective advantages to them. Again, widespread testing is essential to this purpose. However, different from the simpler RT-qPCR-based diagnostic assay to detect SARS-CoV2 RNA in suspected patients, detecting the occurrence and circulation of genetic variants is a much more complex task. To date, the monitoring of viral variants’ arising and circulation is done almost exclusively through NGS [31–33]. That can be a huge constraint for the global monitoring of SARS-CoV2 mutants, as equipment and trained personal to perform the technique and analyze the results are not broadly available, especially in developing and under-developed countries where the epidemics soar out of control.

Two recent and somewhat opposed examples illustrate the importance of early detection of SARS-CoV2 variants to avoid their uncontrolled spread. The B.1.617 variant, usually called Indian variant, was first reported in late 2020 at the Maharashtra Indian state [34], and today has spread to more than 40 countries. After the first identification of B.1.617 variant, three subtypes with slightly different set of mutations have been described through a massive effort using NGS to track the spread and evolution of this new variant [35]. The early detection of this VOC harboring a worrisome set of mutations previously related to increased virus infectivity and partial escape from antibody neutralization has helped to create an important state of preparedness abroad. In UK, coordinated efforts to monitor the spread of the Indian variants through massive acquisition of NGS data has helped scientists and sanitary authorities to model the new mutants’ spread within the country. This network has been able to determine, for example, that the B.1.617.2 subtype has outcompeted the other two Indian B.1.617 subtypes and now seems to be replacing the former prevalent B.1.1.7 variant [35].

The second example comes for the P.1 VOC arising in Brazil in November 2020. The P.1 harbors a plethora of lineage-defining mutations, including some particularly worrisome modification in the S protein that have been related to increased transmissibility (K417T, E484K, and N501Y) and neutralization antibodies’ partial escape (E484K) [9,36]. The rise of this new VOC has been pointed out as one of the pivotal reasons for the recent and overwhelming wave of SARS-CoV2 cases in the city of Manaus, the capital of the Amazon State, leading to a full collapse of the city’s health system [10]. This shocking wave of infections and deaths took place despite the fact that most of the city’s population was thought to be immune after a massive first wave of COVID-19 hit the city in March/April 2020 [36]. Still, the new P.1 variant circulated undetected for months until it was described in four Japanese travelers returning from Manaus to Japan [37]. Only then, tipped by that finding, scientists were able to detected the virus circulating in Manaus through networking NGS efforts [10,36]. By then, the P.1 variant had already spread from Manaus and quickly became the prevalent variant circulating in many other Brazilian regions [38].

If the P.1 variant had been detected earlier, it is possible to speculate that the second large COVID-19 wave hitting Brazil could have been at least partially attenuated. And it is also possible to speculate that constraints in doing massive NGS throughout the country can be partially responsible for such failure. Sanger nucleotide sequencing is much easier to execute and analyze and largely available in different research laboratories than NGS. When tailored to focus in genomic regions bearing signature mutations, the sequence length limitation of the Sanger sequencing strategy ceases to be a problem and become an advantage, as a much larger number of samples can be sequenced in a shorter period of time. The data present here showed that the method was able to accurately detect the appearance of the P.1 VOC in two of the largest Brazilian cities and was also able to unveil the strong capacity of the P.1 variant to disseminate and quickly replace the P.2 variant, which was prevalent lineage in both cities. Although beyond the scope of this study, a better understanding of the molecular mechanism behind the increased dissemination capacity as well as the implications of the spreading of distinct variants regarding vaccination and other methods for the control of the pandemic are certainly necessary. Further efforts towards mitigation of new COVID-19 waves may include studies that rely on a broad implementation of Sanger sequencing genotyping strategy such as the one described here.

## Data Availability

Sequences generated using the Sanger method are available in GenBank:
identifiers MZ410314 to MZ410541; and sequences generated using the Ion Torrent Platform are
available in GISAID data bank: identifiers EPI_ISL_2551523 to EPI_ISL_2551536,
and EPI_ISL_2627737, EPI_ISL_2627738.

## Data availability

Sequences generated using the Sanger method are available in GenBank [39] with identifiers **MZ410314** to **MZ410541**, and sequences generated using the Ion Torrent Platform are available in GISAID data bank [40] with identifiers **EPI_ISL_2551523** to **EPI_ISL_2551536**, and **EPI_ISL_2627737, EPI_ISL_2627738**.

## Acknowledgments

We thank Dr. Renato S. Aguiar, Dr. Betânia P. Drumond and Dr. Jônatas S. Abrahão for technical discussions, Dr. Unaí Tupinambás, Dr. Claudia R. L. Alves, Dr. Elaine L. Machado and MSc Murilo S. Costa for setting up the PBH-UPA cohort. We also thank Dr. Pedro Augusto Alves and Carolina P. Rocha por sequencing reactions performed at the Plataforma de Sequenciamento Capilar do Centro de Pesquisa Rene Rachou and Igor Pereira Godinho for administrative support. We also thanks Dr. Milena De Paulis from University Hospital at São Paulo University (HU-USP); Dr. Flavia Jaqueline Almeida from Santa Casa da Misericórdia Hospital (SCMH); Dr. Andressa Simões Aguiar from Hospital São Luiz Gonzaga (HSLG), Infant Hospital Candido Fontoura (IHCF) and Geriatric Hospital Dom Pedro; Dr. Luciana Becker Mau from Pediatric Hospital Menino Jesus (PHMJ) and Dr. Carolina Sucupira from Infant Hospital Darcy Vargas.

## Financial support

This work was supported by the Brazilian Ministry of Science, Technology and Innovation (MCTI) through the “Rede Virus” initiative and its many individual projects: Sub-rede Corona-ômica, sub-rede Diagnóstico, sub-rede Laboratórios de Campanha. The research leading to these results also received funding from the Secretaria de Ensino Superior do Ministério da Educação [grant number SEI 23072.211119/2020-10], from FAPEMIG (Fundação de Amparo à Pesquisa do Estado de Minas Gerais), Finep (Financiadora de Estudos e Projetos), from Coordenação de Aperfeiçoamento de Pessoal de Ensino Superior (CAPES), from CNPq (Conselho Nacional de Desenvolvimento Científico e Tecnológico), from FAPESP (Fundação de Amparo à Pesquisa do Estado de São Paulo) [Project 2020/06409-1] as well as from the Sociedade Beneficiente Israelita Brasileira Hospital Albert Einstein and Associação Brasileira de gestão em projetos. GPS, EGD, BLT, FGF, SMRT, APF are recipients of CNPq fellowships.

---

**Supplementary table.**
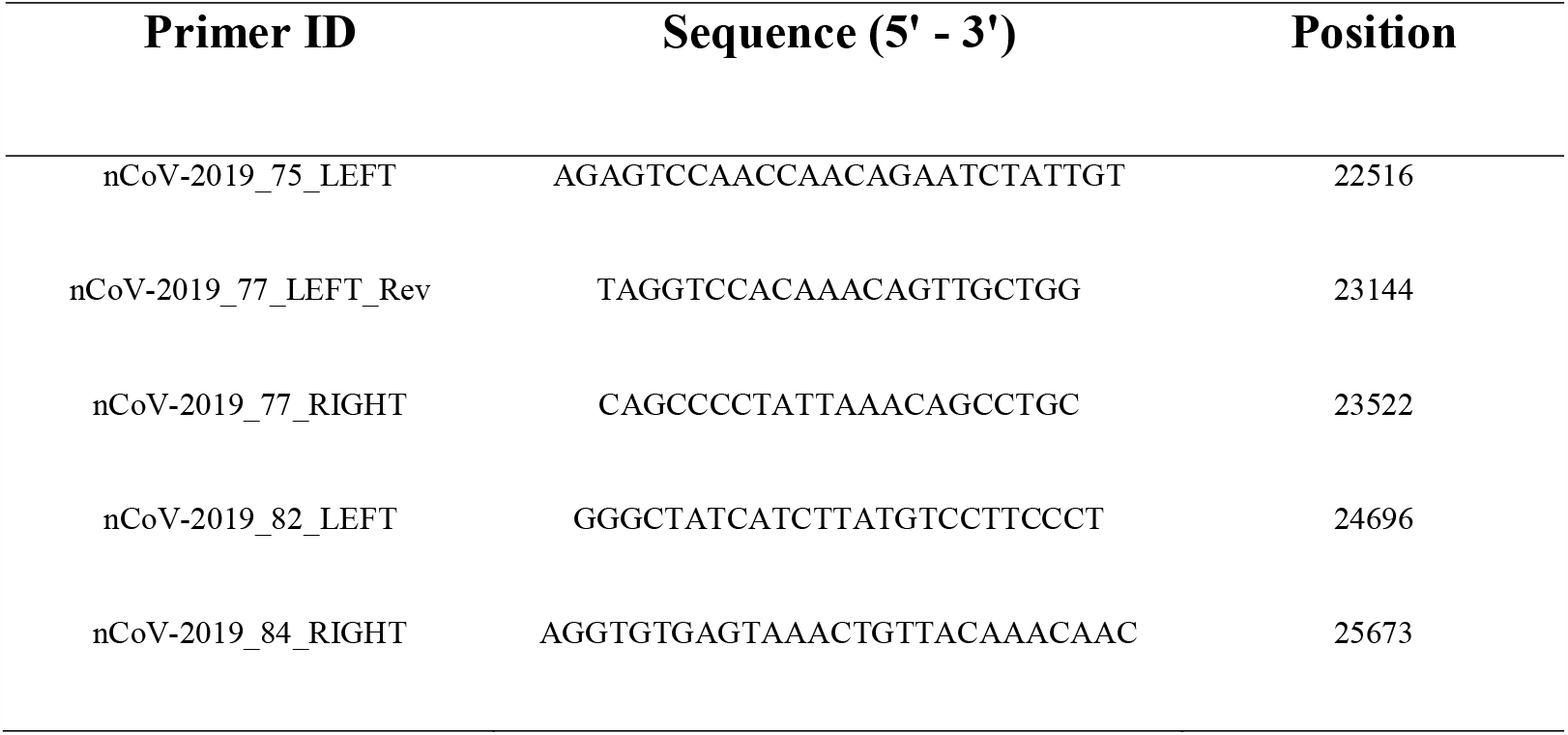
Primers used for PCR amplification and Sanger sequencing. In the table are present the identifiers and sequences of the primers (Primer ID, Sequence (5’ – 3’)), the position of the hybridization of each primer on virus genome (Position). The nCoV-2019_75_LEFT, nCoV-2019_77_RIGHT and nCoV 2019_77_LEFT_Rev primers were used for obtention of 1006 bp fragment sequence and the nCoV-2019_82_LEFT and nCoV-2019_84_RIGHT primers were used for 969 bp fragment.

## Notes

### Competing Interest Statement

The authors have declared no competing interest.

### Author Declarations

Part of study involving samples from Sao Paulo city was approved by Instituto de Ciencias Biomedicas ethical committee. CAAE number: 32712620.9.0000.5467. The study involving samples from Belo Horizonte was approved by the UFMG Ethics Committee, CAAE- 35074720.3.0000.5149.

